# Impact of Bilateral Transmuscular Quadratus Lumborum Block Using Different Doses of Dexmedetomidine as an Adjuvant Combined with Ropivacaine for Postoperative Analgesia in Laparoscopic Myomectomy

**DOI:** 10.1101/2020.08.19.20177725

**Authors:** Yue Li, Le Zhang, Jing Jiao, Xinhua Yu, Shaoqiang Huang

## Abstract

**Introduction:** The optimal dosage of dexmedetomidine (DEX) as an adjuvant combined with ropivacaine for postoperative analgesia in laparoscopic myomectomy is still controversial. The main aim of this prospective clinical trial was to evaluate the analgesic effect and relevant adverse effects of different doses of dexmedetomidine applied locally for ropivacaine-induced bilateral transmuscular quadratus lumborum block (TQLB).

**Methods:** TQLB was conducted using different doses of dexmedetomidine per side (R group: control group; DEX1 group: 0.1μg/kg; DEX2 group: 0.3μg/kg; DEX3 group: 0.5μg/kg). Numeric rating scales (NRS) of pain score, heart rate (HR) and blood pressure (BP) were assessed at different time points after performing TQLB. Dosage of additional analgesics via patient-controlled analgesia pump, recovery time for anal exsufflation, hospital stay and clinical events such as nausea, vomiting and pruritus were also compared among groups.

**Results:** A total of 200 patients underwent laparoscopic myomectomy were enrolled in this study and divided into 4 groups (R, DEX1, DEX2 and DEX3 group) randomly, 50 for each group. Compared with R group, NRS pain score was significantly lower in DEX2 group and DEX3group (p<0.001) 10 minutes after TQLB. Twenty minutes after TQLB, statistical significance of NRS pain score appeared between DEX1 group and R group (p<0.05) and between DEX2, 3 group and DEX1 group (p<0.001). In comparison to R group and DEX1 group, NRS pain score in DEX2 and DEX3 group were lower within 24 hours postoperatively (p<0.05). HR in DEX2 group and DEX3 group was significantly lower than DEX1 group and R group (p<0.05). 2 patients in DEX2 and DEX3 group were developed severe low HR of which HR was below 50bpm. Even 6 hours after surgery, patients in DEX3 group still remained severe low HR. Patients in DEX2 group and DEX3 group needed fewer PCA bolus deliveries than patients in R group and DEX1 group (p<0.001). Satisfaction score of postoperative analgesia was higher in DEX2 group and DEX3 group in comparison to DEX1 group and R group (p<0.001).group and DEX3 group in comparison to DEX1 group and R group (p<0.001).

**Conclusion:** 0.3μg/kg per side of dexmedetomidine as an adjuvant for TQLB was recommended to effectively relieve postoperative pain after laparoscopic myomectomy.

**Trial registration:** Chinese Clinical Trial Registry with registration number ChiCTR1900028509. Date of registration: 24 November 2019. http://www.chictr.org.cn/edit.aspx?pid=42635=htm=4

## INTRODUCTION

Laparoscopic myomectomy is a common procedure in gynecological surgery. Compared with open abdominal surgery, it has less damage, less bleeding and shorter hospital stay^[1]^, which are accordant with the concept of Enhanced Recovery after Surgery (ERAS). For rapid recovery, adequate and suitable postoperative analgesia is also important. Several studies have shown that transversus abdominis plane (TAP) block contributes to less postoperative nausea and vomiting (PONV) ^[2, 3]^ and earlier removal of urinary catheter ^[4]^. Similarly, quadratus lumborum block is an effective technique, provides more potent and prolonged postoperative analgesia in comparison to the TAP block ^[5]^, and is helpful for reducing opioid use as well as postoperative rehabilitation. With the growing interest in the use of transmuscular quadratus lumborum block (TQLB) for postoperative analgesia of laparoscopic myomectomy, it is imperative to find out an optimal medication scheme to improve the effect of analgesia.

Dexmedetomidine (DEX) has been demonstrated to be an effective adjuvant to local anesthetics for peripheral nerve block. Local application of dexmedetomidine plays an important role in shortening onset time ^[6, 8]^, extending duration of analgesia for nerve block without neurotoxicity^[6–8, 9]^ and reducing the dosage of local anesthetics. However, little is known about the dose-response effects of perineural dexmedetomidine. Clinical dose of locally applied dexmedetomidine varies among studies of different kinds of perineural nerve block, most of which were greater than or equal to 1μg/kg. Since higher dosage of dexmedetomidine may reduce heart rates and cause bradycardia due to its sympatholytic effect, a suitable dosage of dexmedetomidine which is both safe and effective for TQLB is desirable.

Therefore, our primary aim was to evaluate the analgesic effect of transmuscular quadratus lumborum block using different doses of dexmedetomidine as an adjuvant combined with ropivacaine in laparoscopic myomectomy. Our secondary aim was to compare adverse effects and outcomes of patients among different medication schemes.

## METHODS

### Study design and participants

This prospective randomized controlled trial was approved by the Institutional Review Board and was registered in the Clinical Research Information Service (ChiCTR1900028509).

The study was conducted between December 2019 and July 2020. Female patients who had ASA physical status I and II and underwent laparoscopic myomectomy were included and randomized into one of four groups (R group, DEX1 group, DEX2 group and DEX3 group) (Figure 1). Eligible participants were free of diabetes mellitus, psychosis, shock, herpes zoster, neurodynia and local anesthetic allergy at the baseline. We also excluded drug abusers, opioid addicts and patients who were not able to understand NRS or not able to use PCA pump. Informed consent was obtained from all participants before enrollment. Those who were converted to open laparotomy, loss of follow-up, had pathological malignant and intraoperative complications such as anaphylactic shock and massive haemorrhage were also excluded in the final analysis.

**Figure 1.**
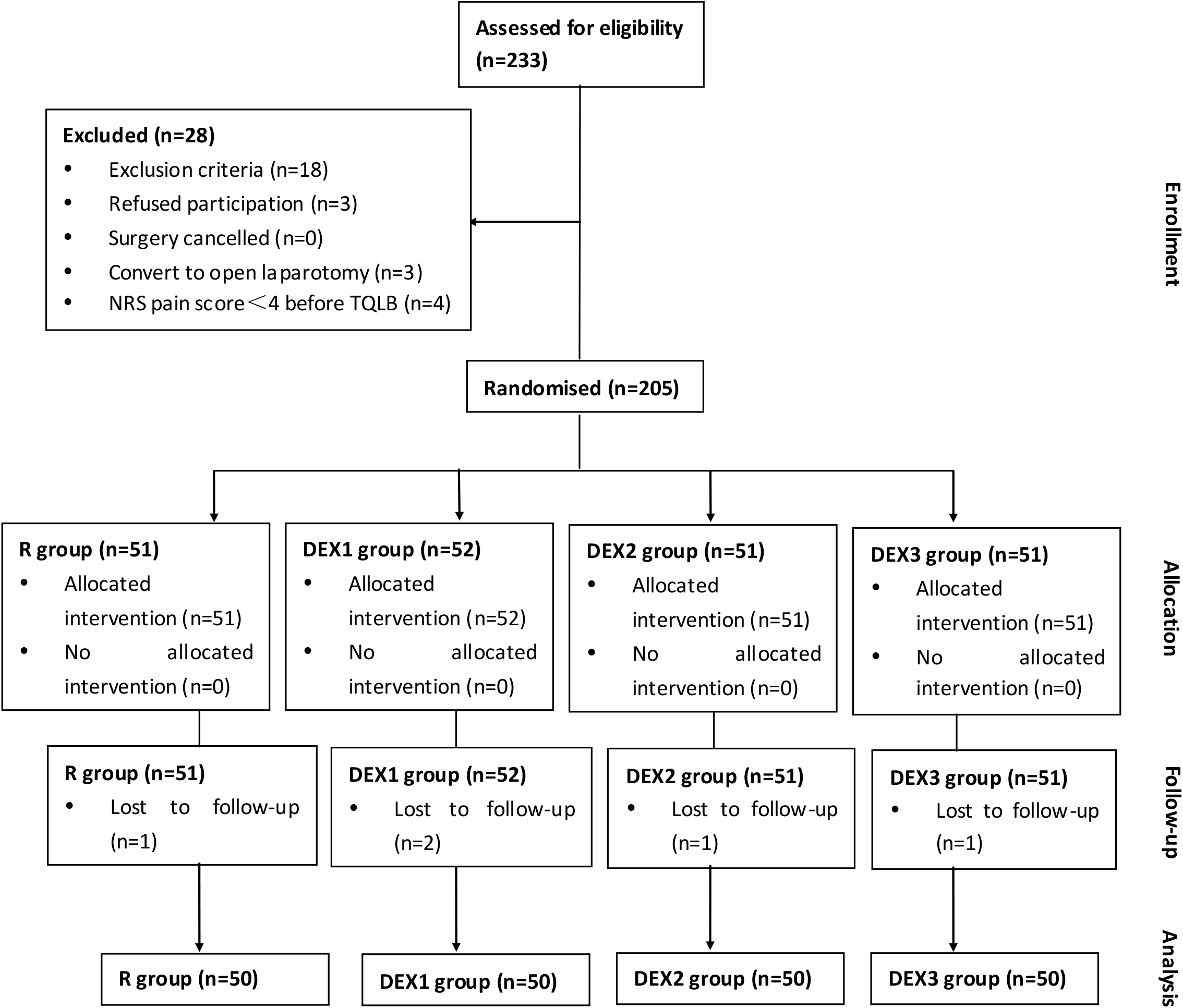
Consort flow chart of recruitment. R, ropivacaine; DEX, dexmedetomidine

Participants were randomly assigned to one of four groups in a 1:1:1:1 ratio using a computer-generated randomization number sequence. Preparation of local anesthetic was performed by a nurse who was not involved in the study and blinded to the patient assignment status. Patients and anesthesiologists were all blinded to group assignment.

### Conduct of anesthesia and postoperative analgesia

In accordance with our routine protocol, general anesthesia was induced with sufentanil 0.5 μg/kg, dexamethasone 5mg, rocuronium 8mg/kg and propofol 4μg/ml TCI. After tracheal intubation, anesthesia was maintained with desflurance 5%–6% and remifentanil 8–10μg/kg/h. Rocuronium was injected intermittently to achieve adequate muscle relaxation. NSAIDs and other analgesics such as oxycodone and tramadol were not allowed to use intraoperatively. After operation, patients were extubated and transported into postanesthesia care unit (PACU). Routine monitoring was performed in PACU using non-invasive blood pressure, electrocardiography and pulse oxygen saturation.

Before conducting ultrasound-guided TQLB, we used numeric rating scales (NRS) to evaluate the intensity of pain. Then patients were positioned lateral decubitus with their knees bent. A low-frequency convex probe was placed with a transverse orientation lateral to the L3 spinous process. Quadratus lumborum, L3 spinous process, psoas major muscle and erector spinae muscle were clearly visible in the image. The needle was inserted between the quadrates lumborum and psoas major muscle using the ‘in-plane’ ultrasound-guided technique. For dexmedetomidine groups, a total 20ml of 0.25% ropivacaine and different doses of dexmedetomidine (DEX1 group: 0.1 μg/kg; DEX2 group: 0.3μg/kg; DEX3 group: 0.5μg/kg)was injected per side. And for the control group (R group), ropivacaine was injected without dexmedetomidine. Patients were informed to use the PCA pump when their pain was not under control. The PCA pump was set to deliver a 2μg sufentanil and 2mg nabulphine bolus dose on demand without continuous infusion, with a lock time of 15 minutes.

### Postoperative assessment

In PACU, NRS pain score, blood pressure and heart rate were recorded at 10, 20, 30, 60 minutes after the TQLB procedure to evaluate the onset and effect of analgesia. Likewise, NRS was also used 6, 24 and 48 hours postoperatively to assess the probable duration of analgesia by comparing with the baseline NRS pain score which was evaluated before the conduct of TQLB. The highest sensory blocked level was tested in PACU 30 minutes after TQLB was performed.

For adverse effects, heart rate (HR) below 60 bpm was defined as low HR, while HR below 50 bpm was defined as severe low HR. Blood pressure (BP) below 90/60 mmHg was called low BP. All those data were recorded in PACU and in the follow-up period. For patients with severe low HR, 0.5 or 1mg of atropine was used to increase the heart rate.

Sedation was assessed using the Ramsay score (1= irritable; 2= quiet, cooperative and with good orientation; 3= drowsy, but still responsive to commands; 4= light sleep, but still active when tapping the forehead; 5= sleep, and dull to the forehead tapping stimulation; 6= deep sleep, having no response to the forehead tapping stimulation). Patients who scored more than 3 were recorded as an adverse effect of dexmedetomidine. In addition, satisfaction was assessed using 5-point rating scale (1= Not at all Satisfied; 2= Partly Satisfied; 3= Satisfied; 4= More than Satisfied; 5= Very Satisfied). Dosage of additional analgesics via PCA pump in 24 hours, exhaust time, hospital stay and clinical features like nausea, vomiting and pruritus were recorded to help to reflect the effect of analgesia and ERAS to a certain extent.

### Statistical analysis

In the pilot study that compared the changes of postoperative NRS pain scores between DEX and R groups, 60 patients were randomly divided into 4 groups (R, DEX1, DEX2 and DEX3 group), with 15 patients for each group. The dosage of dexmedetomidine in each group was the same as that in our present study. The estimated mean NRS pain score in the R group was 3.6 points with a standard deviation (SD) of 1.24. The estimated mean NRS pain scores in the DEX1, DEX2 and DEX groups were 2.6 (1.12), 2.1 (1.22), and 1.9 (1.25), respectively. Thus, based on one-way ANOVA with 4 groups, a two-sided type I error (α) of 0.05, and a power of 80%, the required sample size per group was 44. Taking an anticipated drop-out rate of 20% into consideration, we planned to recruit a total of 212 patients with 53 patients in each group.

NRS pain score, HR, BP and data associated with outcomes among groups were presented as mean ± standard deviation (SD). The median highest sensory blocked level in each group and the number of PCA bolus were presented as median and interquartile range (IQR). For parametric data, the four groups were compared using one-way ANOVA, and for non-parametric data, Kruskal-Wallis tests were used. Since the main comparisons were DEX groups versus R and DEX2 and DEX3 groups versus DEX1, we did not explicitly adjust for multiple comparisons. Chi-squared test was used for comparing the incidence of PONV. Statistical significance was set at P< 0.05. Statistical analysis was performed using SPSS 20.0 (SPSS, Inc., Chicago, IL, USA) and GraphPad Prism 5.01 software (GraphPad Inc., CA, USA).

## RESULTS

### Demographic Characteristics and intraoperative variables

Consort flow chart of recruitment was shown in Figure 1. Initially, 233 patients were assessed for eligibility and 28 were excluded because of exclusion criteria (n=18), refused participation (n=3), conversion to open laparotomy (n=3) and NRS pain score <4 before TQLB (n=4). The remaining 205 patients were randomly divided into 4 groups (R, DEX1, DEX2 and DEX3 group). Two patients in the DEX1 group and one patient in each R, DEX2 and DEX3 group were lost to follow-up. The final analysis consisted of a total of 200 patients who underwent laparoscopic myomectomy. As shown in Table 1, there were no differences in patients’ demographic characteristics such as ages, height and weight. American Standards Association (ASA) classes and data of surgery, including blood loss and operation time did not attain statistical significance as well (P > 0.05, Table 1).

**Table 1.** Demographic characteristics and intraoperative variables (mean±SD)

|  | R | DEX1 | DEX2 | DEX3 | P-value |
| --- | --- | --- | --- | --- | --- |
| Age, y | 40.3±10.7 | 40.5±11.9 | 40.5±9.7 | 40.2±9.9 | 0.968 |
| Height, cm | 160.2±5.6 | 161.2±4.1 | 162.2±5.4 | 161.0±5.3 | 0.368 |
| Weight, kg | 56.3±7.1 | 59.4±8.0 | 58.3±9.6 | 56.5±6.8 | 0.228 |
| Blood loss, ml | 104.7±123.4 | 104.5±111.9 | 106.4±132.4 | 120.9±204.8 | 0.945 |
| Operation time, min | 89.3±37.9 | 78.3±34.9 | 75.1±30.7 | 80.7±24.9 | 0.258 |

R= ropivacaine; DEX=dexmedetomidine.

### Analgesic effect

Thirty minutes after TQLB, the median highest sensory blocked level in each group was T7. No statistical difference was found among groups (R group: IQR, 7.0–8.0; Dex1 group: IQR, 7.0–8.0; DEX2 group: IQR, 6.0–8.0; DEX3 group: IQR, 7.0–8.0; p=0.096). The baseline NRS pain scores assessed before TQLB was performed were not statistically different (p=0.079). During the follow up, compared with R group, NRS pain score was significantly lower in the DEX2 group and DEX3 group (p<0.001) 10 minutes after TQLB. Twenty minutes after TQLB, statistical significance of NRS pain score emerged in comparisons between the DEX1 group and the R group (p<0.05) and between the DEX2,DEX3 group and the DEX1 group (p<0.001) (Table 2).

**Table 2.** Numeric rating scales (NRS) score after TQLB (mean±SD).

|  | R | DEX1 | DEX2 | DEX3 |
| --- | --- | --- | --- | --- |
| Baseline | 5.2±1.3 | 4.9±1.1 | 4.7±1.4 | 5.3±1.5 |
| 10min | 4.4±1.5 | 3.8±1.0 | 3.0±1.4*<sup>#</sup> | 3.26±1.3* |
| 20min | 3.6±1.1 | 2.9±0.9* | 2.1±1.1*<sup>#</sup> | 2.3±1.1*<sup>#</sup> |
| 30min | 3.3±1.2 | 2.5±1.0* | 1.8±0.9*<sup>#</sup> | 1.78±0.8*<sup>#</sup> |
| 60min | 3.1±1.1 | 2.4±0.9* | 1.6±0.8*<sup>#</sup> | 1.62±0.8*<sup>#</sup> |
| 6h(rest) | 2.7±1.0 | 2.2±1.0 | 1.3±0.7*<sup>#</sup> | 1.12±0.8*<sup>#</sup> |
| 6h(movement) | 3.6±1.5 | 3±1.1 | 2.2±1.0*<sup>#</sup> | 2±1.0*<sup>#</sup> |
| 24h(rest) | 2.8±1.3 | 2.3±0.9 | 1.5±1.0*<sup>#</sup> | 1.5±1.1*<sup>#</sup> |
| 24h(movement) | 3.8±1.5 | 3.3±1.2 | 2.7±1.3* | 2.6±1.3*<sup>#</sup> |
| 48h(rest) | 1.2±0.7 | 1.2±0.7 | 1.1±0.9 | 1.0±0.9 |
| 48h(movement) | 2.4±1.0 | 2.5±1.3 | 2.1±1.1 | 2.0±1.1 |

\*p < 0.05 was considered statistically significant compared with the R group. #p < 0.05 was considered statistically significant compared with the DEX1 group. R= ropivacaine; DEX=dexmedetomidine; h=hour.

NRS pain score was also evaluated 6, 24 and 48 hours after surgery, at rest and with movement respectively. NRS pain score was significantly lower in the DEX2 and DEX3 groups in comparison to the R and DEX1 groups within 24 hours postoperatively (p<0.05). No statistical significance was found between the DEX2 group and the DEX3 group at any time point (Table 2).

### Postoperative adverse reactions

During the period in PACU, there was a downward trend of HR in all 4 groups. Before TQLB, no significant difference was found among four groups (p=0.956). Ten minutes after TQLB, HR in the DEX2 and DEX3 group was significantly lower than that of the DEX1 and R group (Figure 2).

**Figure 2.**
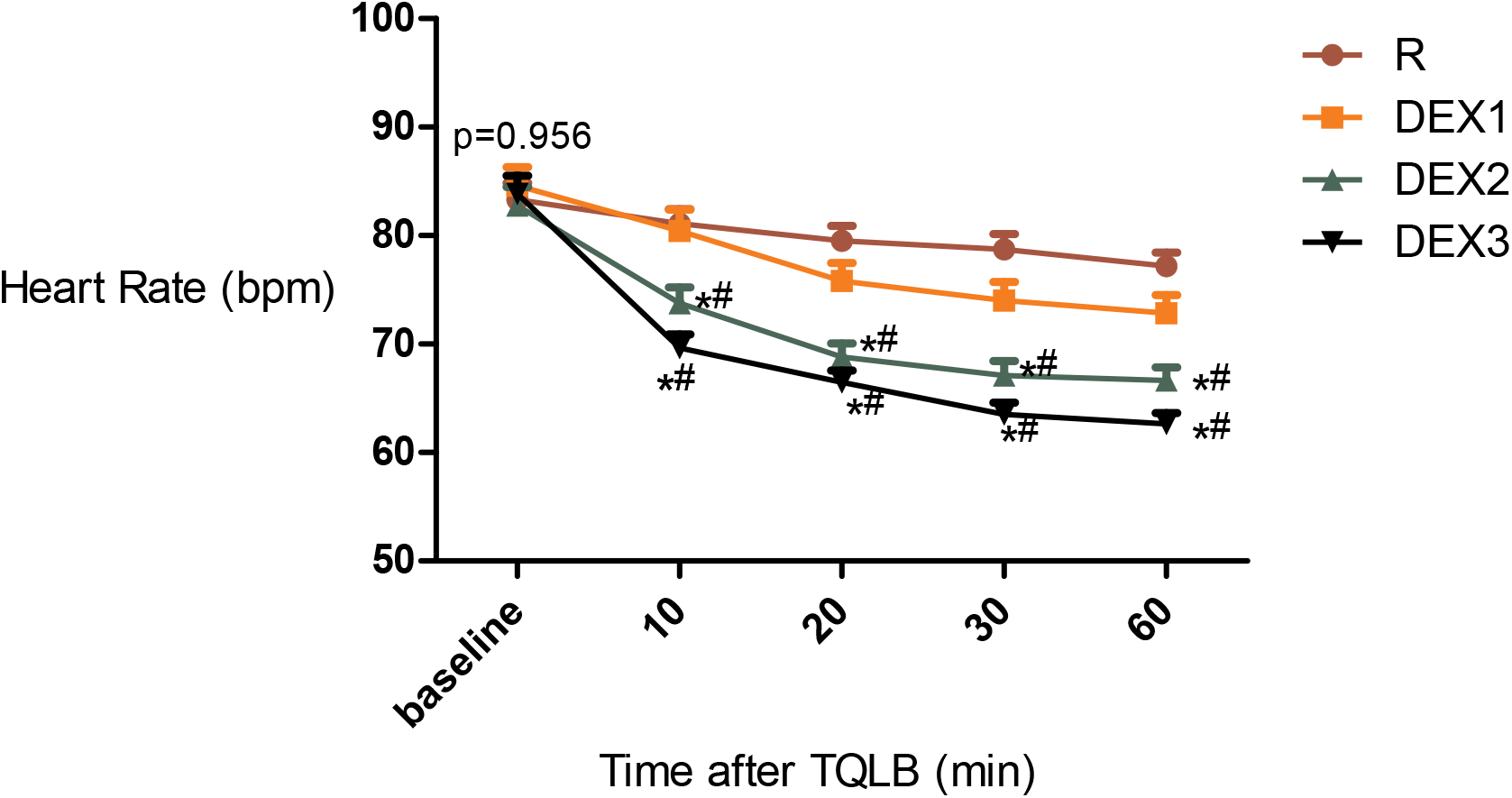
The variation of heart rate at different time points after the performance of TQLB. (*p < 0.05 was considered statistically significant compared with the R group; #p < 0.05 was considered statistically significant compared with the DEX1 group; baseline = time immediately after the performance of TQLB)

There were 8 patients in the DEX1 group, 17 in the DEX2 group and 22 in the DEX3 group that were found having low HR (HR<60bpm) in PACU. Two patients in each DEX2 and DEX3 group were developed severe low HR (HR<50bpm). Even in 6 hours after surgery, one patient in the DEX2 group and three patients in the DEX3 group were diagnosed of low HR, and two patients remained severe low HR in the DEX3 group. There was no statistical significance in the incidence of nausea (p=0.288) and the number of vomiting (p=0.652) among 4 groups (Table 3). One patient in the DEX2 group and three patients in the DEX3 group were found having low blood pressure in PACU after TQLB. Six hours after surgery, there were 2 patients in the DEX3 group whose blood pressure was still below 90/60 mmHg. Symptom of pruritus was not found in all patients. Some patients in the DEX2 and DEX3 groups reported drowsy symptoms after TQLB, but the Ramsay scores among four groups were all below 3 points.

**Table 3.** The incidence of nausea and the number of vomiting after surgery

|  | R |  | DEX1 |  | DEX2 |  | DEX3 |  | P-value |
| --- | --- | --- | --- | --- | --- | --- | --- | --- | --- |
|  | n | % | n | % | n | % | n | % |  |
| Nausea | 14 | 28 | 7 | 14 | 9 | 18 | 8 | 16 | 0.288 |
| Vomiting |  |  |  |  |  |  |  |  |  |
| 0 | 39 | 78 | 38 | 76 | 40 | 80 | 40 | 80 | 0.652 |
| 1 | 5 | 10 | 6 | 12 | 4 | 8 | 5 | 10 |  |
| 2 | 3 | 6 | 5 | 10 | 1 | 2 | 1 | 2 |  |
| 3 | 1 | 2 | 1 | 2 | 3 | 6 | 1 | 2 |  |
| 4 | 2 | 4 | 0 | 0 | 2 | 4 | 3 | 6 |  |

R= ropivacaine; DEX=dexmedetomidine.

### Other outcomes

Patients in the DEX2 and DEX3 groups needed significantly fewer PCA bolus deliveries than patients in the R and DEX1 groups (p <0.001). Satisfaction score of postoperative analgesia achieved on the third postoperative day was significantly higher in the DEX2 and DEX3 groups comparing to the DEX1 and R groups (p <0.0001). The recovery time for anal exsuffflation (p=0.096) and the average postoperative hospital stay (p=0.152) among groups did not differ (Table 4).

**Table 4.**
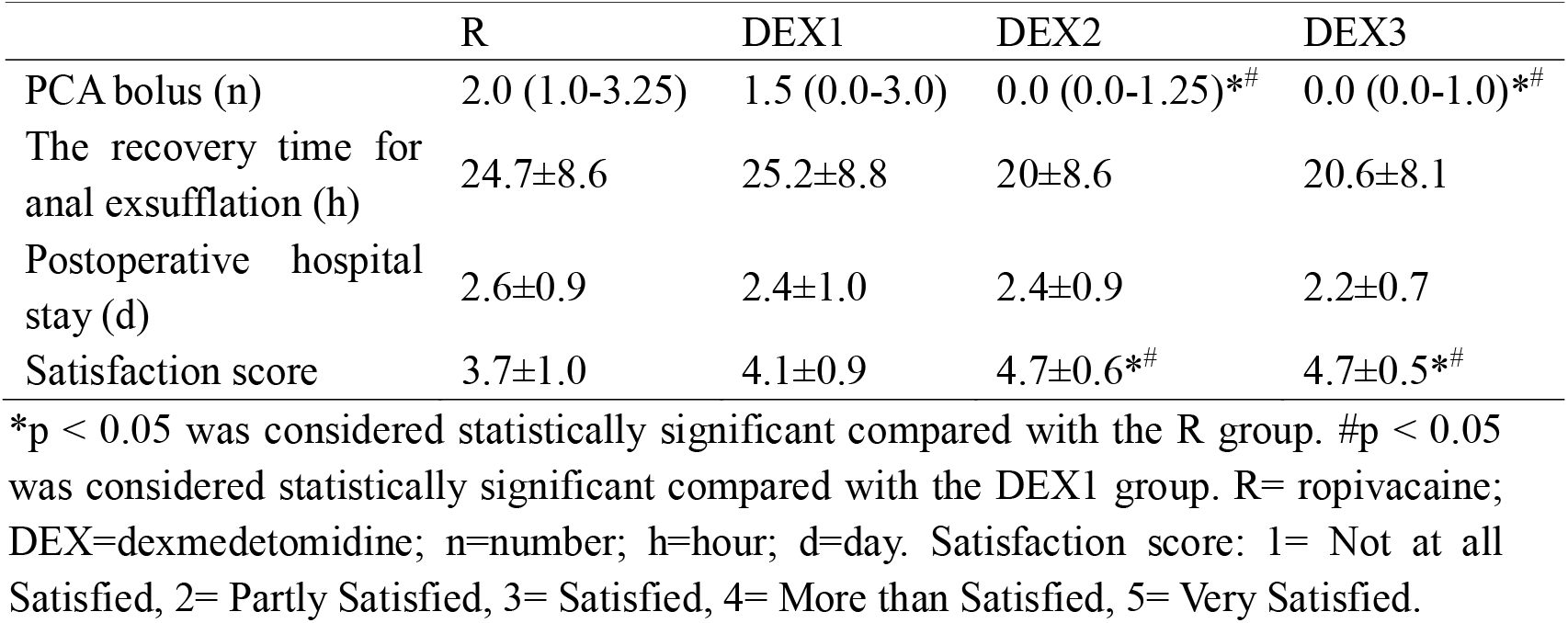
Other outcomes and satisfaction of postoperative analgesia. Data are median (IQR) or mean±SD

## DISCUSSION

In this study, we found that local administration of dexmedetomidine greater than or equal to 0.3μg/kg per side for TQLB (DEX2, DEX3) improved postoperative analgesia effect for up to 24 hours. It has been reported in many studies that the addition of dexmedetomidine to local anesthetics for peripheral nerve block shortens the onset and prolongs the duration of analgesia. Since the estimated minimum significant variation in NRS pain score is 1.4 ^[10]^, substantial change in NRS pain score of pain occurred 20 min after the performance of TQLB in the R and DEX1 groups in our study. In contrast, in the DEX2 and DEX3 groups, significant clinical change in NRS pain score occurred 10 min after TQLB. In other words, the onset of analgesia was shortened when the dosage of dexmedetomidine as an adjuvant to ropivacaine-induced TQLB was no less than 0.3μg/kg per side.

The mechanism of local injected dexmedetomidine for postoperative pain control is complicated. Anti-inflammatory effect and vasoconstriction ^[11, 12]^at the injected site were be possible mechanisms.α2 adrenoceptor agonists, including dexmedetomidine, also have an inhibitory effect on Na+ channels in neurons, which may explain its enhanced effect on analgesia to some extent.

Suitable dosage of dexmedetomidine varies among different kinds of peripheral nerve block. Jian Yu et al in 2018^[13]^ showed that 1.5μg/kg DEX exhibited a superior effect in improving ROP induced lumbar plexus sciatic nerve block. A study published in 2008^[14]^ recommended 2μg/kg as the clinical dose of perineural administration of dexmedetomidine. In 2017, Asku et al^[15]^ found that local administration of 1μg/kg of dexmedetomidine reduced the demand of bupivacaine to half. Vallapu et al^[16]^ in 2018 showed that the duration of analgesia was longer when 1μg/kg of dexmedetomidine was added as an adjuvant to local anesthetic agent in scalp block. Some of these studies neglected the occurrence of adverse effects and the other reported cases of over-sedation, bradycardia and dry mouth in high-dosage group. In our previous clinical practice, one patient was reported severe bradycardia (HR 43 bpm) while the dosage of dexmedetomidine was 0.6μg/kg per side (a total of 1.2μg/kg). Hence, we chose relatively low doses (0.1, 0.3 and 0.5μg/kg per side) to observe their effectiveness. In our study, it was obvious that less than 0.3μg/kg dexmedetomidine per side did not effectively enhance the analgesia action of ropivacaine-induced TQLB, and 0.3μg/kg dexmedetomidine per side provided the same analgesic effect compared with 0.5μg/kg.HR in all groups decreased after the implementation of bilateral transmuscular quadratus lumborum block (Figure 2), which was possibly associated with the control of postoperative pain. However, HR in the DEX2 and DEX3 groups was significant lower than that in the R and DEX1 groups at each time point in PACU after the performance of TQLB. This was probably attributed to the inhibitory effect of dexmedetomidine on sympathetic nerve tension ^[17]^.

Caution should be taken when a total of 0.6μg/kg or more dexmedetomidine was used as an additive to TQLB in patients with bradycardia since severe low HR (HR<50bpm) occurred in both DEX2 and DEX3 group in PACU. Severe bradycardia in DEX3 group had even lasted for up to 6 hours postoperatively. Thus, for TQLB in laparoscopic myomectomy, 0.3μg/kg of dexmedetomidine per side was safer than 0.5μg/kg of dexmedetomidine, which provided the same analgesic effect.

Although the number of PCA bolus was significantly higher in the R and DEX1 groups in contrast to the DEX2 and DEX3 groups, the incidence of nausea and vomiting had no statistical significance among groups in our study. This might possibly because that the PCA bolus dosage (2μg sufentanil and 2mg nabulphine) was not enough to result in obvious nausea and vomiting.

For other outcomes, there was no significant difference in hospital stay and the recovery time for anal exsufflation. However, satisfaction score in the DEX2 and DEX3 groups were higher than in the R and DEX1 groups. Satisfaction with analgesia is an important outcome in many clinical studies ^[18]^. In our study, local administration of no less than 0.3μg/kg per side of dexmedetomidine for TQLB led to mild sedation, which helped patients to rest and further increased patients’ satisfaction with analgesia.

There were several limitations in the present study. First, it was difficult to observe the duration analgesia among groups because postoperative pain of laparoscopic myomectomy was relieved spontaneously in two days after surgery. Second, the optimal dosage of dexmedetomidine in our study was achieved by parallelized randomized controlled trials, which divided participants into 3 groups of different doses of dexmedetomidine, instead of sequential trial because it was difficult to define whether the intervention was effective or not. 0.3μg/kg per side of dexmedetomidine for TQLB was what we found a relatively suitable dosage compared with 0.1μg/kg and 0.5μg/kg. In addition, in order to avoid interference factors that might affect the actual analgesic effect of TQLB, NSAIDs was not allowed to use perioperatively, which more or less weaken the postoperative analgesia in comparison to our routine protocol. Theoretically, the use of NSAIDs can reduce the amount of other analgesic drugs to some extent, so the dose of dexmedetomidine more than 0.3μg/kg per side is more unnecessary in combination with NSAIDs. And the impact of locally applied dexmedetimidine combined with ropivacaine for TQLB when NSAIDs are used in combination needs further research.

## Conclusion

An optimal dosage of dexmedetomidine for ropivacaine-induced TQLB should provide maximum analgesia with relatively less hemodynamic changes ^[19–21]^. In our study,0.3μg/kg per side of dexmedetomidine as an adjuvant for TQLB was recommended to effectively relieve postoperative pain after laparoscopic myomectomy. Patients with bradycardia had better make prudent use of more than 0.3μg/kg dexmedetomidine per side for TQLB.

## Data Availability

The datasets generated and analyzed during the current study are available from the corresponding author on reasonable request.

## List of abbreviations

(TQLB): transmuscular quadratus lumborum block
(NRS): Numeric rating scales
(DEX): dexmedetomidine
(ERAS): Enhanced Recovery after Surgery
(TAP): transversus abdominis plane
(PONV): postoperative nausea and vomiting
(PACU): postanesthesia care unit
(ASA): American Standards Association
(TCI): Target controll infusion
(PACU): Postanesthesia care unit
(HR): Heart rate
(BP): Blood pressure
(SD): Standard deviation
(IQR): Interquartile range

## DECLARATIONS AND ACKNOWLEDGMENTS

### Ethical approval and consent to participate

Ethical approval was given by the institutional ethics committee (IRB of Obstetric and Gynecology Hospital of Fudan University) and registered at Chinese Clinical Trial Registry with registration number ChiCTR1900028509.

Written informed consent of participation is obtained from all participants.

### Consent to publish

Not applicable.

## Authors’ Contributions

LY data recording, analysis, paper writing and part of clinical studies. ZL part of clinical studies. HSQ experimental design. JJ equipment providing. All authors read and approved the final version of the manuscript.

## Acknowledgments

Thanks are due to Lin Tian for assistance with the experiments and to Liqun Yang for valuable discussion.

